# Adverse Events During a 12-month Multi-Site and Dose-Response Aerobic Exercise Intervention

**DOI:** 10.1101/2024.09.10.24313352

**Authors:** Eric D. Vidoni, George Grove, Amanda N. Szabo-Reed, Mickeal N. Key, Haiqing Huang, Jeffrey M. Burns, Charles H. Hillman, John M. Jakicic, Chaeryon Kang, Arthur F. Kramer, Edward McAuley, Lu Wan, Tera Hawes, Sydney S. White, Kirk I. Erickson

## Abstract

**Purpose:** This study aimed to assess the incidence of adverse events (AE) in older adults participating in a year-long exercise intervention, investigating potential dose-response relationships between exercise intensity and AE frequency, and identifying demographic factors associated with AE risk.

**Methods:** A total of 648 older adults were randomized into one of three exercise groups: low-intensity stretching and toning (S&T), 150 minutes of aerobic exercise per week (150Ex), or 225 minutes of aerobic exercise per week (225Ex). Adverse events were tracked during the intervention, with event rates calculated based on participant adherence and time in the study. Generalized linear models were employed to compare AE incidence across groups. Post hoc comparisons were used to calculate incidence rate ratios (IRRs) for AE between groups, adjusting for multiple comparisons.

**Results:** Overall, 306 AE were reported, with 44% related to the intervention. No significant dose-response relationship was observed for all-cause AE between groups. However, intervention-related AE were more frequent in the aerobic exercise groups. Participants in the 150Ex group had a 77% higher rate of intervention-related AE compared to the S&T group, and the 225Ex group had an 88% higher rate. Higher adherence was associated with fewer all-cause AE, and greater comorbid burden was associated with more AE.

**Conclusions:** While aerobic exercise increased the risk of intervention-related AE, the overall risk remained low. Higher adherence to the exercise regimen was associated with fewer AE. These findings suggest aerobic exercise is generally safe in older adults, with the benefits outweighing the risks.

## INTRODUCTION

Exercise is among the most important health behaviors across the lifespan(1). Exercise is beneficial for cardiovascular,(2) pulmonary,(3) musculoskeletal,(4, 5) and endocrine health.(6) However, exercise is also associated with injuries with more than 8 million medically attended sport and recreation injuries occurring in the United States each year.(7) This incidence does not account for the many more non-medically attended injuries that might prevent or modify exercise behaviors and resolve with self-treatment. Indeed, there is little descriptive evidence regarding the adverse, unintended health consequences of exercise beyond these broad estimates, especially amongst older adults.

Randomized controlled trials (RCT), with rigorous adverse event (AE), or “medical harms”, reporting are an important potential source of information on exercise-related injuries. AE, typically defined as any untoward medical occurrence (8) are a required component of the Consolidated Standards of Reporting Trials (CONSORT) framework. Standardized reporting of physiologic system involvement, severity, and relatedness, along with other trial data such as demographics may allow for a richer description of exercise risks and benefits in a specific population and minimize the incidence of AE in future exercise trials. The rigor of modern safety monitoring allows for examining the frequency of exercise-related injury and the relationship to other functional factors. However, these important trial experiences are frequently condensed in favor of main outcomes and offered with limited context or investigation.

Recognizing the potential value of analyzing the frequency of adverse health changes, both related to and independent of exercise, we explored the AE incidence in the Investigating Gains in Neurocognition in an Intervention Trial of Exercise (IGNITE) study (NCT02875301). IGNITE was a multi-site exercise RCT in 648 older adults that was designed to examine the effects of aerobic exercise on cognitive and brain outcomes in late adulthood. IGNITE provided a structured and rigorous foundation to explore several important questions about safety and AE frequency amongst older adults participating in a 12-month exercise regimen: (1) Is there a dose-response relationship between exercise and AE? (2) What is the relative frequency of exercise-related AE to non-related AE? (3) Are there demographic profiles that are more likely to experience exercise-related AE? We hypothesized that the frequency of AE and exercise would vary in relation to the dose of exercise prescribed, with greater doses related to a higher frequency of AE. Further we predicted that individuals with greater comorbid and socioeconomic burden would be at a greater risk for experiencing exercise-related AE.

## METHODS

IGNITE enrolled 648 older adults without cognitive impairment or major morbidity into a 1-year exercise trial to measure changes in cognition, brain health, and other outcomes under a previously described protocol.(9) The three sites used a common electronic data capture system, REDCap,(10) allowing for standardized capture of study data including AE. Briefly, participants were block randomized accounting for age and sex into a low intensity stretching and toning control group (S&T), 150 minutes (150Ex) or 225 minutes (250Ex) of moderate intensity aerobic exercise (40-60% heart rate reserve), in equal ratio. Participants exercised with supervision of a personal trainer or similar certified professional up to 3 times per week with two additional unsupervised sessions 2 times per week. Blood collection and testing for cognitive function, physical function, social and medical history, cardiorespiratory fitness (peak oxygen consumption during a graded exercise test, VO_2_peak), mental health, brain health, and other outcomes were performed at baseline and 52 weeks, with additional cognitive testing and blood collection at 26 weeks. Germane to this analysis, state-level Area Deprivation Index (11) based on participants’ residence at consent, and the Cumulative Illness Rating Scale – Geriatric (CIRS-G) (12) were collected during baseline testing. Exercise adherence was measured as percentage of the prescribed duration completed as reported by the participant on weekly exercise logs. Primary and secondary outcomes will be reported in other manuscripts. The study was approved by the Institutional Review Board of Pittsburgh in adherence to the Declaration of Helsinki, with reliance agreements from other sites. All participants provided written, informed consent.

Study staff were trained to perform AE inquiry at regular “check-in” intervals approximately every two weeks and at every point of person-to-person communication (e.g. unscheduled gym visits and testing sessions). The typical participant would be questioned regarding health changes approximately 33 times by study staff. Trainers were also trained to report medical history changes to the study team. Any reported change in the medical condition of the participant was recorded by study staff at each site as an AE on a standard form that included classifying the physiologic system involved, the relatedness to the study (“Not Related” or “Possibly or Probably Related”), and designation as a Serious Adverse Event (SAE) reportable to our Data and Safety Monitoring Board and sponsor. SAE included hospitalization, life threatening conditions, and death.

As part of IGNITE standard procedure, pre-existing conditions “discovered” through research testing procedures were characterized. These incidental findings included identification of electrocardiographic evidence of heart disease and abnormal neuroimaging, for example. Typically, these incidental findings required additional clinical review, which delayed enrollment and added to study cost and participant burden. Information on the incidence of these pre-existing conditions in the literature is sparse. Projections of these incidental findings are valuable to those prospectively planning trials and are reported here, separately from AE.

We compared the incidence of adverse events across intervention groups using generalized linear models with a Poisson distribution.(13) The models included the intervention group as the independent variable and the number of adverse events as the dependent variable, with an offset for the log-transformed observation period (from consent to the end of testing or participation). A random intercept for each participant was included. Time in trial varied by participant due to reasons such as early withdrawal and COVID-19 lockdown intervention extensions. We assessed model fit by checking for overdispersion and zero-inflation and alternatively fitted a negative binomial and/or zero-inflation corrected regression model if indicated. Following that, we conducted likelihood ratio tests between the full model, including the independent variable, and a reduced model with only the intercept and offset to evaluate the significance of the independent variable as a predictor of adverse event incidence rates. For *post hoc* analysis, we computed estimated marginal means for each category of the independent variable, followed by pairwise comparisons with Tukey’s adjustment for multiple comparisons to control the family-wise error rate across all pairwise tests. The resulting incident rate ratios (IRR) from the model reflect the incidence of adverse events per year between categories of the independent variable.

For demographic comparisons, we divided by the sample median for age (<u>> vs <</u> 70y), CIRS-G Total Score (≥4 vs. less), Area Deprivation Index (ADI, a measure of socioeconomic disadvantage, 4^th^ state-adjusted decile and above vs. below) and cardiorespiratory fitness (VO_2_peak ≥ 20ml/kg/min vs. below), Body Mass Index (BMI) greater than 30 (a clinically relevant cutoff), and sex (female vs. male). Descriptive tables include total AE counts, average counts per person, and average counts per person-year, aiming to provide insights into AE frequency for future studies. All statistical analyses were conducted in R (ver. 4.4.0) using the ‘base’, ‘emmeans’, ‘glmmTMB’, and ‘MASS’ packages.

## RESULTS

### Adverse Events Overview

The study team recorded 144 incidental findings and 20 adverse events (AE) during pre-intervention and post-intervention assessment sessions and 306 AE during the intervention, from the 648 individuals randomized into the IGNITE study. Demographics and AE totals are detailed in Table 1.

**Table 1:** Demographics of IGNITE Sample.

|  | Intervention Group |  |  |  |
| --- | --- | --- | --- | --- |
|  | Overall<br>N = 648 | S&T<br>N = 210 | 150Ex<br>N = 217 | 225Ex<br>N = 221 |
| Age, yrs | 69.8 (3.7) | 69.9 (3.7) | 69.7 (3.8) | 69.8 (3.8) |
| Female, n | 461 / 648<br>(71.1%) | 150 / 210<br>(71.4%) | 154 / 217<br>(71.0%) | 157 / 221<br>(71.0%) |
| Education | 16.3 (2.2) | 16.3 (2.2) | 16.4 (2.3) | 16.2 (2.2) |
| CIRS-G Total Score | 3.3 (2.4) | 3.2 (2.5) | 3.4 (2.4) | 3.3 (2.4) |
| Cardiorespiratory fitness, ml/kg/min | 21.7 (5.1) | 21.5 (5.1) | 21.7 (4.8) | 21.8 (5.3) |
| Area Deprivation Index, decile | 3.6 (2.7) | 3.6 (2.7) | 3.6 (2.7) | 3.6 (2.8) |
| Intervention Adherence, % prescribed | 78 (38) | 80 (37) | 82 (37) | 74 (39) |
| Average AE Count Per Person | 0.47 (0.79) | 0.45 (0.81) | 0.47 (0.76) | 0.49 (0.81) |
| Average Intervention-Related AE<br>Count Per Person | 0.21 (0.50) | 0.13 (0.38) | 0.24 (0.56) | 0.25 (0.54) |
| Average AE Count Per Person-Year | 0.54 (1.54) | 0.53 (1.94) | 0.52 (1.16) | 0.56 (1.44) |
| Average Intervention Related AE<br>Count Per Person-Year | 0.21 (0.66) | 0.16 (0.72) | 0.24 (0.72) | 0.23 (0.52) |
| <sup>1</sup>Mean (SD); n / N (%) |  |  |  |  |

### Incidental Findings and Adverse Events Outside the Intervention Period

About 1/5^th^ of our participants (n = 134) who were randomized were flagged for an incidental finding that required further evaluation. In total, the 144 incidental findings from these participants were mostly cardiovascular (n = 82) or neurological (n = 53), with 9 other miscellaneous findings. The findings predominantly included electrocardiogram abnormalities evident during exercise testing and MRI abnormalities noted during a mandatory radiology read. Participants also experienced 20 AE during the testing periods. These were coincidental in timing and unrelated to study activities and consisted mostly of upper respiratory infections and musculoskeletal injuries sustained in activities of daily living. Three SAE due to hospitalizations were reported prior to randomization. None of the SAE were related to testing procedures.

### Intervention and Adverse Event Patterns

Because our primary intent for this manuscript was to characterize incidence of AE associated with an exercise intervention, the remaining analyses focus on post-randomization AE. In total, 219 participants reported 306 all-cause AE during their intervention participation (those defined as either related or unrelated to the intervention); 134 were adjudicated to be related to the intervention, and 172 were not related to the intervention. Our primary hypothesis that AE frequency would increase with greater prescribed exercise doses was not supported, as there was no significant dose-response observed for all-cause AE (Χ^2^(2) = 0.23, p = 0.89). Participants experienced 0.47 (SD 0.79) AE on average during their time in the study. Counts for all-cause AE can be found in Table 2.

**Table 2:**
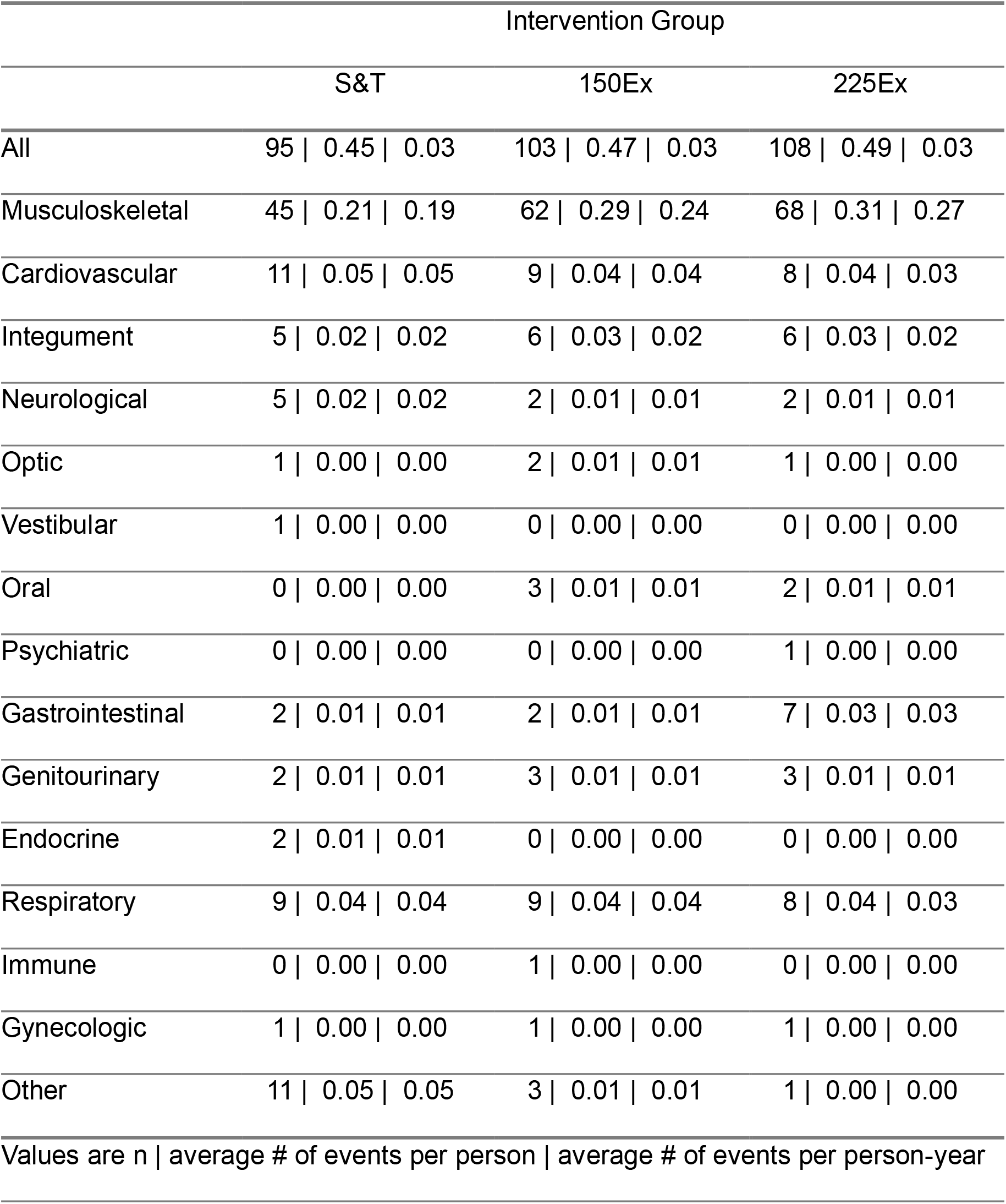
All-cause AE by System During the Intervention Period.

|  | Intervention Group |  |  |
| --- | --- | --- | --- |
|  | S&T | 150Ex | 225Ex |
| All | 95 \| 0.45 \| 0.03 | 103 \| 0.47 \| 0.03 | 108 \| 0.49 \| 0.03 |
| Musculoskeletal | 45 \| 0.21 \| 0.19 | 62 \| 0.29 \| 0.24 | 68 \| 0.31 \| 0.27 |
| Cardiovascular | 11 \| 0.05 \| 0.05 | 9 \| 0.04 \| 0.04 | 8 \| 0.04 \| 0.03 |
| Integument | 5 \| 0.02 \| 0.02 | 6 \| 0.03 \| 0.02 | 6 \| 0.03 \| 0.02 |
| Neurological | 5 \| 0.02 \| 0.02 | 2 \| 0.01 \| 0.01 | 2 \| 0.01 \| 0.01 |
| Optic | 1 \| 0.00 \| 0.00 | 2 \| 0.01 \| 0.01 | 1 \| 0.00 \| 0.00 |
| Vestibular | 1 \| 0.00 \| 0.00 | 0 \| 0.00 \| 0.00 | 0 \| 0.00 \| 0.00 |
| Oral | 0 \| 0.00 \| 0.00 | 3 \| 0.01 \| 0.01 | 2 \| 0.01 \| 0.01 |
| Psychiatric | 0 \| 0.00 \| 0.00 | 0 \| 0.00 \| 0.00 | 1 \| 0.00 \| 0.00 |
| Gastrointestinal | 2 \| 0.01 \| 0.01 | 2 \| 0.01 \| 0.01 | 7 \| 0.03 \| 0.03 |
| Genitourinary | 2 \| 0.01 \| 0.01 | 3 \| 0.01 \| 0.01 | 3 \| 0.01 \| 0.01 |
| Endocrine | 2 \| 0.01 \| 0.01 | 0 \| 0.00 \| 0.00 | 0 \| 0.00 \| 0.00 |
| Respiratory | 9 \| 0.04 \| 0.04 | 9 \| 0.04 \| 0.04 | 8 \| 0.04 \| 0.03 |
| Immune | 0 \| 0.00 \| 0.00 | 1 \| 0.00 \| 0.00 | 0 \| 0.00 \| 0.00 |
| Gynecologic | 1 \| 0.00 \| 0.00 | 1 \| 0.00 \| 0.00 | 1 \| 0.00 \| 0.00 |
| Other | 11 \| 0.05 \| 0.05 | 3 \| 0.01 \| 0.01 | 1 \| 0.00 \| 0.00 |
| Values are n \| average # of events per person \| average # of events per person-year |  |  |  |

When specifically looking at AE judged to be intervention-related, we found that incidence rate ratios (IRR) differed between intervention groups (Χ^2^(2) = 7.05, p = 0.03), but not in a dose-dependent manner. Participants in the 150Ex group trended toward a 77% higher incidence of AE compared to the S&T group (IRR = 1.77, 95% CI [0.96, 3.25], p = 0.07). Participants in the 225Ex group had an 88% higher incidence of AE compared to the S&T group (IRR = 1.88, 95% CI [1.03, 3.43], p = 0.038). There was no significant difference in the incidence of AE between the 150Ex and 225Ex groups (IRR = 1.05, 95% CI [0.63, 1.78], p = 0.96). Participants experienced 0.21 (SD 0.66) injuries or other health changes related to exercise per year. Counts for intervention-related AE can be found in Table 3.

**Table 3:** Intervention-Related AE by System During the Intervention Period.

|  | Intervention Group |  |  |
| --- | --- | --- | --- |
|  | S&T | 150Ex | 225Ex |
| All | 28 \| 0.13 \| 0.01 | 51 \| 0.24 \| 0.01 | 55 \| 0.25 \| 0.01 |
| Musculoskeletal | 22 \| 0.10 \| 0.09 | 47 \| 0.22 \| 0.18 | 47 \| 0.21 \| 0.18 |
| Cardiovascular | 6 \| 0.03 \| 0.02 | 4 \| 0.02 \| 0.02 | 1 \| 0.00 \| 0.00 |
| Integument | 0 \| 0.00 \| 0.00 | 0 \| 0.00 \| 0.00 | 2 \| 0.01 \| 0.01 |
| Neurological | 0 \| 0.00 \| 0.00 | 0 \| 0.00 \| 0.00 | 1 \| 0.00 \| 0.00 |
| Optic | 0 \| 0.00 \| 0.00 | 0 \| 0.00 \| 0.00 | 0 \| 0.00 \| 0.00 |
| Vestibular | 0 \| 0.00 \| 0.00 | 0 \| 0.00 \| 0.00 | 0 \| 0.00 \| 0.00 |
| Oral | 0 \| 0.00 \| 0.00 | 0 \| 0.00 \| 0.00 | 0 \| 0.00 \| 0.00 |
| Psychiatric | 0 \| 0.00 \| 0.00 | 0 \| 0.00 \| 0.00 | 0 \| 0.00 \| 0.00 |
| Gastrointestinal | 0 \| 0.00 \| 0.00 | 0 \| 0.00 \| 0.00 | 1 \| 0.00 \| 0.00 |
| Genitourinary | 0 \| 0.00 \| 0.00 | 0 \| 0.00 \| 0.00 | 1 \| 0.00 \| 0.00 |
| Endocrine | 0 \| 0.00 \| 0.00 | 0 \| 0.00 \| 0.00 | 0 \| 0.00 \| 0.00 |
| Respiratory | 0 \| 0.00 \| 0.00 | 0 \| 0.00 \| 0.00 | 1 \| 0.00 \| 0.00 |
| Immune | 0 \| 0.00 \| 0.00 | 0 \| 0.00 \| 0.00 | 0 \| 0.00 \| 0.00 |
| Gynecologic | 0 \| 0.00 \| 0.00 | 0 \| 0.00 \| 0.00 | 0 \| 0.00 \| 0.00 |
| Other | 0 \| 0.00 \| 0.00 | 0 \| 0.00 \| 0.00 | 1 \| 0.00 \| 0.00 |
| Values are n \| average # of events per person \| average # of events per person-year |  |  |  |

We also assessed AE frequency in the musculoskeletal and cardiovascular systems since these are colloquially associated with exercise injury. There was no significant effect of intervention group on all-cause musculoskeletal AE (Χ^2^(2) = 2.81, p = 0.25). But much like overall AE, intervention group was a significant factor in musculoskeletal AE related to the intervention (Χ^2^(2) = 8.40, p = 0.02). Participants in the 150Ex group had a 105% higher incidence of intervention-related musculoskeletal AE compared to the S&T group (IRR = 2.05, 95% CI [1.07, 3.95], p = 0.03). Similarly, participants in the 225Ex group had a 103% higher incidence of intervention-related musculoskeletal AE compared to the S&T group (IRR = 2.03, 95% CI [1.05, 3.91], p = 0.03). There was no significant difference in the incidence of musculoskeletal AE between the 150Ex and 225Ex groups (IRR = 0.98, 95% CI [0.57, 1.70], p = 0.99). There was no difference in related or all-cause cardiac AE between groups (p > 0.22). There was also no group difference in the incidence rate of non-intervention related AE (Χ^2^(2) = 1.96, p = 0.37).

Finally, we assessed whether exercise prescription adherence was related to AE frequency after accounting for time on intervention. Adherence was significantly associated with fewer all-cause AE (IRR = 0.53, 95% CI: [0.29,0.97], p = 0.048). Participants who were fully adherent experienced approximately 47% fewer adverse events. And when considering only intervention-related AE, adherence was not significantly related to AE frequency (IRR = 0.78, 95% CI [0.27, 2.23], p = 0.653).

### Adverse Events by Demographics

Contrary to our secondary hypotheses, age, sex, socioeconomic disadvantage, BMI, and baseline cardiorespiratory fitness were unrelated to AE incidence during intervention (p ≥ 0.08). However, participants with higher comorbidity (CIRS-G Total score at or above he median) had a 40% higher incidence rate of AE compared to those with lower comorbidity burden (Χ^2^(1) = 6.00, p = 0.014, IRR = 1.40, 95% CI [1.07, 1.83], p = 0.01), and this was especially true of intervention-related AE. Participants with higher comorbidity had a 70% higher incidence rate of AE compared to those with lower comorbidity (IRR = 1.70, 95% CI [1.16, 2.50], p < 0.01).

### Serious Adverse Events and Mortality

During the intervention, 24 participants experienced 27 SAE.

Group differences were noted in incidence (Χ^2^(2) = 8.62, p = 0.013). This was driven exclusively by participants in the 150Ex group having 82.1% lower incidence of SAE compared to those in the S&T group (IRR = 0.18, 95% CI [0.04, 0.84], p = 0.03). Three SAE were related to the intervention, one in each intervention group. The related SAE were sudden cardiac death (225 Min), chest pain and hospitalization during exercise (S&T), severe spinal stenosis and hospitalization (150 Min). Additionally, one individual (S&T) was in a motor vehicle accident resulting in death.

## DISCUSSION

Our analysis of AE history in the IGNITE trial represents a novel and important, in-depth look at exercise safety in a large multi-site RCT among relatively healthy older adults. We sought to describe the relative frequency of AE in the context of medical and social history profiles linked to AE in older adults who exercise in a manner consistent with public health recommendations. A key question was whether those mode or duration of exercise impacted AE frequency.

IGNITE offers an opportunity to benchmark AE incidence for partially supervised older adults in exercise programs. Our data suggests that clinical trial staff should expect about 1 medical or health event every 2 years. If engaged in an exercise program targeting a minutes per week and intensity range within public health recommendations, the average adult (65-80 yrs) will experience 0.2 medical or health events related to that exercise program per year, or about 1 injury per 5 exercise years. This low frequency of injury in active older adults should be interpreted as encouraging. Our AE rates are similar to previous reports that about 25% of adults generally engaged in general physical activity will experience an exercise-related injury each year.(14)

Our primary hypothesis of a dose-response relationship between exercise and AE frequency was not supported by the data. There was no difference in the number of total AE per year between the S&T, 150Ex, and 225Ex intervention groups. However, we did find that the aerobic exercise groups experienced more AE deemed “intervention-related”. Our observation of a similar rate of total AE and non-dose-dependence of intervention-related AE across aerobic exercise groups raises several potential interpretations. It is possible that raters experience perception or expectation bias when considering a person in an “active” aerobic exercise group versus the group considered to be a placebo control (S&T). Alternatively, participants in the aerobic exercise groups may be inclined to report AE more frequently. A third possibility is that aerobic exercise conveys some protective benefit against incidental AE unrelated to exercise, such as respiratory infections, which were non-significantly more frequent in the S&T group (see Table 2).

Consistent with conventional logic, we found that AE frequency was inversely related with adherence to the prescribed amount of exercise over the study, even after accounting for time on intervention. That is, people who were sick or injured less, performed more of their full exercise prescription. Of particular interest, however, was that this relationship only held for all-cause AE. Intervention-related AE were not related to adherence. Possible explanations for this include that the greater does of exercise was not sufficient to convey greater exercise-related risk. As seen in Table 1, the 225Ex group was not quite as adherence as the other groups. Alternatively, it is possible that the lack of relationship between intervention-related AE and adherence in these 3 groups, is driven largely by the low-risk nature of the stretching and toning intervention and moderate intensity exercise, both of which are considered safe for almost all individuals with age-appropriate cardiovascular and musculoskeletal function.

In general, we did not find evidence that people with certain demographic characteristics were more likely to report an AE. This included those with a higher BMI, lower cardiorespiratory fitness, age, sex, and those who lived in more resource deprived areas. This is inconsistent with the general understanding of the health risks of being overweight, negative social determinants of health, and poor physical function.(15-18), and could be attributed to a bias of inclusion that is common in many clinical trials.(19, 20) IGNITE did employ a comprehensive list of inclusion and exclusion criteria designed primarily to exclude cognitive impairment from any source. Thus, it is possible that only individuals more resilient to negative health and environmental challenges were selected. However, we found that AE were significantly more frequent in those with greater comorbid burden, which is more in line with common understanding of health risks. Though unreported in our results, we did not identify an interaction of comorbidity and intervention group to suggest that greater intervention duration is of concern for those with greater comorbidity (p = 0.18). Future research should examine whether specific comorbid conditions drive exercise-related AE more than others.

Despite significant rigor, importance, and novelty, there are limitations to this analysis. The IGNITE cohort represents a cognitively unimpaired sample of older adults and a likely-more health resilient population. Analyses such as ours conducted on populations with significant acute or chronic conditions would add to our understanding of exercise safety in the broader population. Additionally, participants received exercise clearance from a licensed provider and underwent baseline testing that identified potential cardiopulmonary risks. This likely biased the sample towards better health. Despite these limitations, our analysis has important implications for lifestyle clinical trials and our understanding of exercise safety for public health recommendations. Our estimates do not inform population-wide estimates of safety but do extend our understanding of the relative frequency of AE and injury. These results are beneficial for the community engaging or seeking to engage in exercise, public health professionals advocating and prescribing exercise, and research teams and regulatory bodies wishing to understand risks associated with exercise. We strongly endorse the rigorous and systematic monitoring of AE in clinical research and the adoption of standardized AE reporting.

In conclusion, we found exercise to be a safe intervention in a cognitively unimpaired sample of older adults cleared for community exercise by a clinician. Higher comorbidity may convey a greater risk of experiencing an adverse event. But the amount and type of commonly prescribed exercise doses does not appear to convey markedly greater risk for adverse medical events. And the well-documented positive benefits of aerobic exercise far outweigh the modifiable risks.

## Data Availability

All data produced in the present study are available upon reasonable request to the authors

